# Machine prescription for chronic migraine

**DOI:** 10.1101/2021.11.07.21265816

**Authors:** Anker Stubberud, Robert Gray, Erling Tronvik, Manjit Matharu, Parashkev Nachev

**Author notes:** M Matharu and P Nachev contributed equally as last authors. **Corresponding author:** Parashkev Nachev, High Dimensional Neurology Group, UCL Queen Square Institute of Neurology, London, UK.

## Abstract

Responsive to treatment individually, chronic migraine remains strikingly resistant collectively, incurring the second-highest population burden of disability worldwide. A heterogeneity of responsiveness, requiring prolonged—currently heuristic— individual evaluation of available treatments, may reflect a diversity of causal mechanisms, or the failure to identify the most important, single causal factor. Distinguishing between these possibilities, now possible through the application of complex modelling to large-scale data, is critical to determining the optimal approach to identifying new interventions in migraine and making the best use of existing ones.

Examining a richly phenotyped cohort of 1446 consecutive unselected patients with chronic migraine, here we use causal multitask Gaussian process models to estimate *individual treatment effects* across ten classes of preventatives. Such modelling enables us to quantify the accessibility of heterogeneous responsiveness to high-dimensional modelling, to infer the likely scale of the underlying causal diversity. We calculate the *treatment effects* in the overall population, and the *conditional treatment effects* among those modelled to respond and compare the true response rates between these two groups. Identifying a difference in response rates between the groups supports a diversity of causal mechanisms. Moreover, we propose a data-driven machine prescription policy, estimating the time-to-response when sequentially trialing preventatives by individualized treatment effects and compare it to expert guideline sequences. All model performances are quantified out-of-sample.

We identify significantly higher true response rates among individuals modelled to respond, compared to the overall population (mean difference of 0.034; 95% CI 0.003 to 0.065; p=0.033), supporting significant heterogeneity of responsiveness and diverse causal mechanisms. The machine prescription policy yields an estimated 35% reduction in time-to-response (3.750 months; 95% CI 3.507 to 3.993; p<0.0001) compared with expert guidelines, with no substantive increase in expense per patient.

We conclude that the highly distributed mode of causation in chronic migraine necessitates high-dimensional modelling for optimal management. Machine prescription should be considered an essential clinical decision-support tool in the future management of chronic migraine.

## Introduction

Migraine presents a therapeutic paradox. It is the second most disabling disease worldwide—first in the 15 to 50 age interval—with enormous social and economic impact.^1-4^ Yet it is considered a treatable disease, responsive to a wide array of readily administered, mechanistically diverse interventions.^5,6^ How do we find ourselves losing a war whose individual battles we are seemingly so well-equipped to win?

Two polar possibilities arise, distinguished by migraine’s currently unknown mode of causation. If its cause is *unitary*—there is a necessary and sufficient mechanism common to all patients—the response to current treatments may be variable because their effect is collateral to the critical disease process. Here finding a new, universally effective agent is theoretically possible, and its effect may be proven in an adequately powered randomized trial.

Conversely, if its cause is *distributed*—there is no single mechanism but a wide, heterogeneous field of interacting causal factors—treatment variability may be explained by varying correspondence between the chosen therapeutic agent and the patient’s specific causal field.^7^ Here our task necessarily complicates to identifying not one but a *family* of mechanisms—and therefore modifying agents—and cannot be plausibly solved by any practicable set of conventional trials, for the unknown fraction of a sample responsive to any given treatment cannot be quantified without an overview of the treatment heterogeneity of the population as a whole.

Reality may fall anywhere between these two extremes. But in relying on randomised controlled trials, the currently dominant approach to therapeutic innovation in migraine excludes the second possibility entirely. It is, moreover, radically at odds with the widespread clinical impression of treatment heterogeneity, and the established practice of speculative, heuristic treatment, optimised by individual feedback over many months.^6^

If a presumption is to be made, it is in favour of distributed, not unitary causation. But in the absence of widely applicable methods of studying complex distributed causation, the distinction has been untestable. The recent advent of highly expressive, computationally-assisted mathematical models now allows us to investigate it empirically, and to address two questions of major translational significance.

First, examining an unselected, consecutive, fully-inclusive, richly phenotyped cohort of 1446 patients with chronic migraine, here we quantify the *individualized treatment effects* of major categories of prophylactic treatment, exploiting causal multitask Gaussian processes models of proven power to extract heterogeneous causal effects from high-dimensional observational data.^8^ In the setting of unitary causation, where individual variability to current agents arises incidentally, there should be no marked difference between treatment effects evaluated across the population—*average treatment effects*—and treatment effects evaluated across the subpopulation identified to be susceptible—*conditional average treatment effects*. Conversely, finding such a difference would support the presence of distributed causation, reflecting consistent individual patterns of diverse mechanistic susceptibility.

Second, if we find individual responsiveness to be determinable, the *order* in which candidate agents are sequentially evaluated in a patient could be objectively optimised. Here we compare the theoretical benefit—quantified in time-to-response— of such *machine prescription* against established heuristic treatment policies, contextualised by estimates of treatment cost. If substantial benefit is observed, machine prescription ought to be preferred over the current expert-driven approach to treatment selection.

To assure generalisability, we quantify all effects on out-of-sample test data, unseen by the models in training. Moreover, our focus is on the *general extent* of achievable treatment individuation and its impact, not the precise observed rank of individual treatments, for our questions seek to establish the correct causal framework in chronic migraine, and the best use of existing evidence in guiding treatment while we await further insight into the aetiology of this complex disorder.

## Materials and methods

### Patients, interventions, and outcomes

An unselected, consecutive, retrospective cohort comprising all patients seen by one neurologist with headache expertise (MM) at the secondary and tertiary Headache Centre at the National Hospital for Neurology and Neurosurgery, Queen Square, UK, from May 2007 to September 2019 was examined. The inclusion criteria were a diagnosis of chronic migraine and the availability of a sufficiently complete structured clinical phenotypic record. Participants were not required to strictly fulfil the diagnostic criteria for chronic migraine,^9^ but all exhibited the distinctive features of migraine as a firm diagnosis. Patients with chronic migraine only considered a differential diagnosis or in cases where the diagnosis was unclear were excluded. All patients evaluated at the Headache Centre routinely undergo a structured clinical assessment, including comprehensive detailed phenotyping and documentation of prior medical history. A proportion undergoes further investigations, including imaging as clinically indicated. Modelling incorporating brain imaging is the subject of a subsequent report. The study population characteristics are provided in Table 1.

**Table 1.** Study population demographics. Legend: SD=standard deviation; IQR=inter-quartile range; ICHD-3=international classification of headache disorders 3^rd^ edition

|  | **All patients (n=1446)** |  | **Fulfilling ICHD-3**<br>**criteria for chronic**<br>**migraine (n=1096)** |  |
| --- | --- | --- | --- | --- |
| Gender female, n (%) | 1042/1445 | (72.1) | 834/1096 | (76.1) |
| Age, mean (SD) | 41.4 | (14.6) | 40.8 | (14.7) |
| Headache frequency in days/month, mean (SD) | 25.4 | (7.6) | 26.9 | (5.3) |
| Exacerbation intensity, mean (SD) | 8.2 | (1.3) | 8.2 | (1.2) |
| Laterality: |  |  |  |  |
| Only unilateral, n (%) | 393/1434 | (27.4) | 272/1090 | (25.0) |
| Only bilateral, n (%) | 565/1434 | (39.4) | 405/1090 | (37.2) |
| Unilateral and bilateral, n (%) | 476/1434 | (33.2) | 413/1090 | (37.9) |
| Throbbing headache, n (%) | 796/1394 | (51.1) | 653/1068 | (61.1) |
| Motion sensitivity, n (%) | 1215/1446 | (84.0) | 1016/1096 | (92.7) |
| Nausea and/or vomiting, n (%) | 1079/1446 | (74.6) | 945/1096 | (86.2) |
| Photophobia, n (%) | 1090/1446 | (75.4) | 944/1096 | (86.1) |
| Phonophobia, n (%) | 1133/1446 | (78.4) | 955/1096 | (87.1) |
| Aura, n (%) | 494/1446 | (34.2) | 452/1096 | (41.2) |
| Cranial autonomic symptoms, n (%) | 819/1446 | (56.6) | 680/1096 | (62.0) |
| 1-2, n (%) | 516/1446 | (35.7) | 422/1096 | (38.5) |
| 3-4, n (%) | 219/1446 | (15.1) | 186/1096 | (17.0) |
| >5, n (%) | 84/1446 | (5.8) | 72/1096 | (6.6) |
| Family history of migraine, n (%) | 817/1366 | (59.8) | 671/1054 | (63.7) |
| Total follow-up time, person-days | 690197 | - | 497060 | - |
| Follow-up time in days, median (IQR) | 135 | (0-652) | 119 | (0-561) |
| Migraine prophylactics tried, median (IQR) | 4 | (2-6) | 4 | (2-7) |
| Effective migraine prophylactics, median (IQR) | 1 | (0-1) | 1 | (0-2) |

The interventions modelled in this study were classed by mode of action and included all preventive therapies for which there were at least 100 adequately documented patient trials. The modelled therapeutic classes were onabotulinumtoxinA, flunarizine, candesartan, serotonin noradrenaline reuptake inhibitors, topiramate, tricyclic antidepressants, acupuncture, valproate, betablockers and serotonergic agents (pizotifen and methysergide). Treatment response for a therapeutic class was defined as positive where more than 50% reduction in headache days was observed, and negative otherwise, by any agent within the class, over an evaluation period of at least three months. Headache days were recorded by patients prospectively on a paper headache diary and evaluated by the neurologist. Treatment responses to onabotulinumtoxinA were labelled as effective only if the PREEMPT paradigm was followed, and a treatment effect remained after the second set of injections, to account for the known high placebo response.

A total of 1831 patients were eligible for inclusion. Of those, 131 were excluded owing to diagnostic uncertainty, and a further 269 owing to missingness, leaving 1446 for analysis (Supplementary Figure 1).

### Data acquisition and data management

Data was collected through automated extraction of the Microsoft Word template-based structured clinical record employed by the Headache Centre. Standard natural language processing techniques such as string matching with regular expressions and grammatical decomposition were used. Accuracy against manual extraction from a held-out subset of 60 patients was 90.73%. Note that this processing was performed for service optimization purposes wholly within the clinical digital environment; all subsequent analysis was performed on data from which all identifiers had been removed.

Categorical and continuous variables were converted to a continuous interval scale. All other features were binarized as present or absent. Supplementary Table 1 outlines details on all included features and outcomes. Samples with more than 10% missing data were removed from the dataset. The mean and standard deviation was used for precision and variance estimates in cases of normal distribution; the median and interquartile range was used otherwise. Normality assumptions were based on visual inspection of histograms and the Shapiro-Wilk test for normality. Effect estimates were reported with 95% confidence intervals. The significance level was set to α=0.004 after Bonferroni correction for 14 comparisons (0.05/14=0.004) in the prescriptive modelling, and the conventional 0.05 in the individualized treatment effect modelling.

### Modelling and statistical analysis

#### Individualized treatment effect modelling

We randomly split the dataset into three stratified subsets: training, validation, and test, the last providing a held-out, out-of-sample definitive benchmark of performance. The partitions were kept separate and created with the following ratios: 4:1 training to test and within the training set, 4:1 training to validation (Supplementary Figure 1). Missing data were imputed with a probabilistic principal component analysis imputer based on the training dataset, and the data was scaled.

To model individualized treatment effects, we implemented a causal multitask Gaussian process model.^8^ The model has been validated to be capable of inferring individualized treatment effects from observational data, accounting for a non-random distribution of the treatment factor. The model learns from the high-dimensional array of features (in our case, headache phenotype and comorbidities) to infer treatment effects. Treatment effects may be interpreted as the theoretical difference in response (here defined as ≥50% reduction in headache frequency) when exposed to a treatment versus not exposed to a treatment. In the implementation of the model, two different interventions are compared, and by learning from the training data, the model can predict treatment effects in unseen data at the individual level—i.e., *individualized treatment effects*. The model was trained and optimized using the training and validation subsets. The discounted cumulative gain was used as a scoring metric to evaluate choice of kernel hyperparameters (Supplementary Table 2). The best performing model in the validation set was finally evaluated on the out-of-sample test set.

We made pairwise comparisons between all prophylactic intervention classes giving individualized treatment effects for each intervention compared with each of the others. Each patient’s individualized treatment effect for an intervention class was calculated as the mean of all pairwise effects including that class. Thus, we arrive at a modelled individualized treatment effect for all intervention classes for each patient. We calculate the *average treatment effects*—i.e., the estimated population treatment effect inferred from the Gaussian process model—as the median and interquartile range of individualized treatment effects for each intervention class. We also report the median and interquartile range of the mean of individualized treatment effects across all pairwise comparisons to provide descriptive in-sample and out-of-sample average treatment effect estimates for each intervention class (Supplementary Figure 2).

Next, we defined a conditional subgroup consisting of the patients whose modelled individualized treatment effect was above the median (owing to skewed data). By calculating the average treatment effect similarly as above for the conditional subgroup, we derive the *conditional average treatment effect*—i.e., each intervention class’ average treatment effect among those predicted by the model to respond. We then compare the *conditional true treatment effect* (the true response rate captured from headache diaries in the conditional subgroup) with the *overall true treatment effect* using a one-sample t-test of the differences across all intervention classes and report the mean difference with a 95% confidence interval. Finally, we estimate the validity of the machine prescription by calculating the 10-fold cross-validated accuracy of a logistic regression model using the predicted individualized treatment effects as the independent variable and the true response as the dependent variable.

#### Impact and prescriptive modelling

To model the impact of machine prescription, we implemented the following strategy: The individualized treatment effects were used to rank the preventative therapies from highest to lowest probability of response for each individual patient. From this, we ascertained the proportion of patients having tried in reality their top three predicted treatments.

Further, given a patient and a sequence of agents ordered according to their response probabilities—i.e., individualized treatment effects—from highest to lowest, *p*_*1*_, *p*_*2*,_ *p*_*3*_ … *p*_*10*_, we calculated the probability of arriving at a treatment success after a given number of failed treatments, *delivered in the optimal predicted sequence*, as follows:

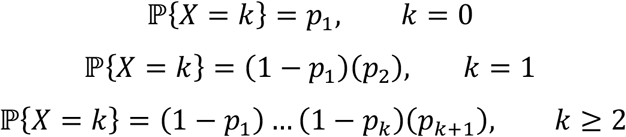

where *X* denotes the number of independent failures before a success (at trial *k+1*). Given treatment success at trial *k+1* we are able to calculate the expected number of months in pain (i.e. months with failed treatments) before completion of a successful treatment trial as

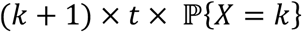

for each patient at *k*=0, *k*=1, *k=*2 … *k*=9. Here *t* equals the necessary time to evaluate a treatment trial which was defined as three months for all treatments except onabotulinumtoxinA which was six months. This gives a population distribution of number of months to completion of a successful treatment trial, allowing us to estimate time-to-response given different sequences of intervention trialling.

We then aimed to evaluate the optimal predicted sequence of intervention trialling to other possible sequences. We compared the population distribution given by the Gaussian process machine prescription to the population distribution given by ranking by different guideline and expert recommendations,^10,11^ following a random sequence, and ordering treatments by increasing costs. We constructed a series of sequences to reflect different available guidelines and expert opinions. Guideline recommendation 1 sequence was constructed by picking three random of the evidence-based oral preventives suggested in at least one guideline (tricyclic antidepressants, serotonin noradrenaline reuptake inhibitors, betablockers, candesartan, topiramate, valproate, and flunarizine) followed by onabotulinumtoxinA.^10^ Guideline recommendation 2 sequence was based on picking two random among betablockers, candesartan, tricyclic antidepressants, and serotonin noradrenaline reuptake inhibitors, followed by one random of topiramate, valproate, and flunarizine, followed by onabotulinumtoxinA.^11^ The National Institute for Health and Care Excellence (NICE) guideline sequence was based on the recommendation of trying each of betablockers, topiramate, and tricyclic antidepressants (order not specified), followed by onabotulinumtoxinA. The expert panel sequence was based on an aggregate of 23 UK headache specialists asked to order the treatments based on a general understanding of efficacy and adverse events (Supplementary Table 3). The random sequence was created from the mean at each timepoint *k*_0_, *k*_1_, *k*_2_ … *k*_9_ of a Monte Carlo simulation with 1000 realizations, i.e. 1000 random sequences. For the machine prescription we also restricted onabotulinumtoxinA to be the fourth trialled treatment to mitigate bias from the difference in evaluation period between onabotulinumtoxinA and other treatments. A two-tailed t-test was used to compare the population distributions of time-to-response, reporting the mean difference with 95% CI.

Using the British National Formulary price tariffs, we derived estimates of individual treatment-related expenses in pound sterling. We then compared the optimal machine prescription sequence of intervention trialling to sequences ordered by treatment costs. Moreover, we reported the difference in expenses defined as the sum of the *n* top predicted trials subtracted from the sum of the *n* actual trials, where *n* is the number of trials. We reported estimates for the lowest and highest available price tariffs (Supplementary Table 4).

We also conducted a sensitivity analysis on sub-strata of severely affected patients vs. less severely affected patients comparing machine prescription to the guideline recommendation 1 sequence. We reiterated the individualized treatment effect and impact analysis on two sub-strata of the population. The first strata consisted of patients with at least 25 headache days/month and a headache intensity of 9 or higher. The second strata consisted of patients with less than 25 headache days/month and headache intensity below 9.

### Ethics

The study was performed under NRES approval by the London-West London & GTAC Research Ethics Committee for the consentless analysis of irrevocably anonymized data.

### Data availability

The raw data required to replicate this study is not availble for public release under the conditions of ethical approval. The code used in this study is available upon reasonable request to the authors.

## Results

### Individualized treatment effects

The out-of-sample modelled *average treatment effects* ranged from 0.44 (interquartile range 0.33 to 0.56) to 0.06 (interquartile range 0.04 to 0.09). The out-of-sample modelled *conditional average treatment effects* ranged from 0.56 (interquartile range 0.48 to 0.65) to 0.09 (interquartile range 0.08 to 0.11). OnabotulinumtoxinA had the largest out-of-sample treatment effects, followed by flunarizine, candesartan, serotonin noradrenaline reuptake inhibitors, topiramate, tricyclic antidepressants, acupuncture, valproate, betablockers, and serotonergic agents.

Out-of-sample comparison of the *true treatment effect* across the population to the *conditional true treatment effect* within the subpopulation of predicted responders for each treatment class showed a mean difference of 0.034 (95% CI 0.003 to 0.065; p=0.033) in favour of the latter (Figure 1). This discrepancy was greatest for flunarizine, serotonergic drugs, and valproate. The accuracy of validating the modelled individualized treatment effects compared to true treatment effects was consistently high (0.731±0.103). Table 2 outlines modelled average treatment effects, modelled conditional average treatment effects, and true treatment effects for all intervention classes.

**Figure 1.**
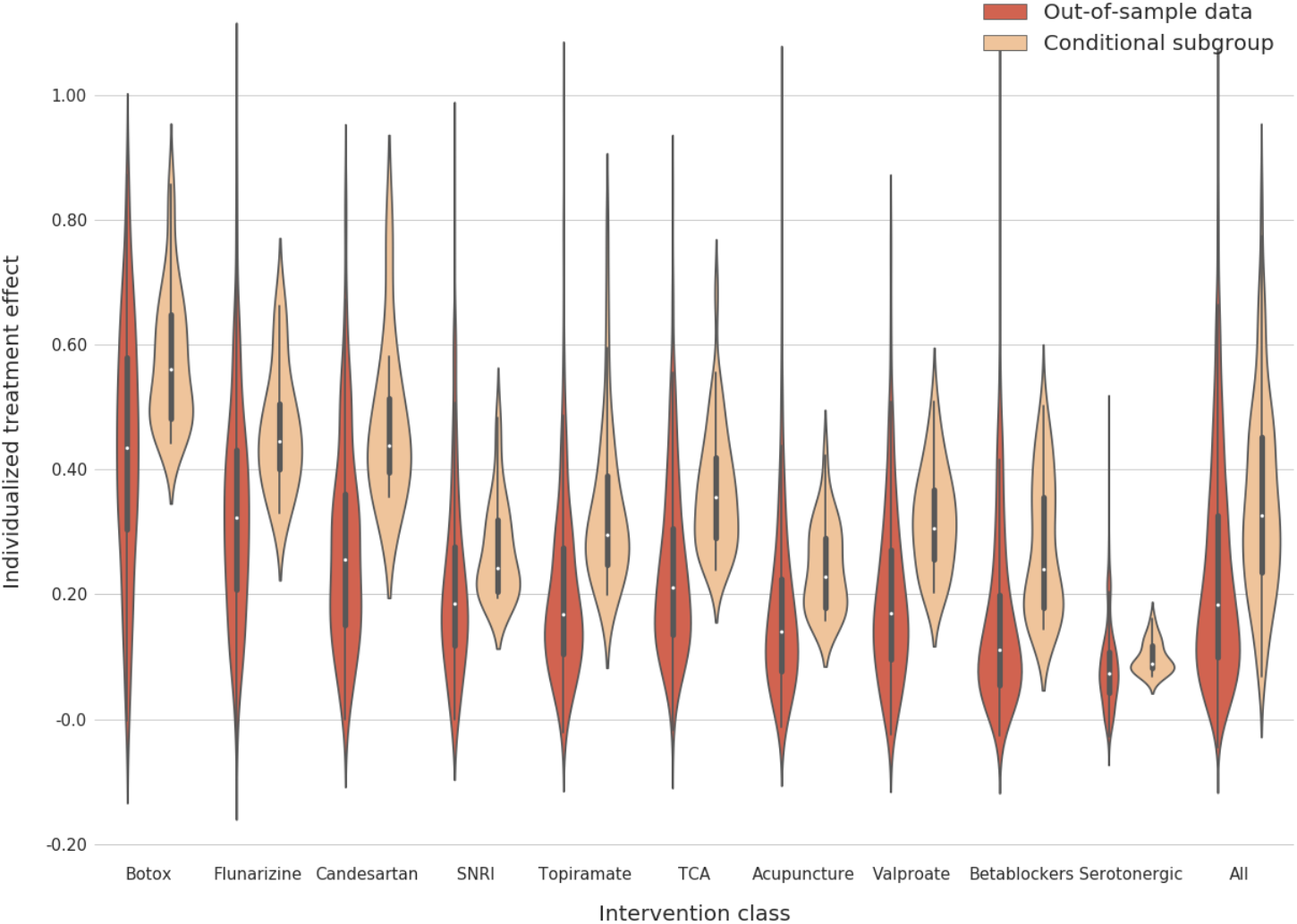
Individualized treatment effects. Violinplot representing modelled individualized treatment effects for overall out-of-sample data (red), and the conditional subgroup predicted to respond (orange). The two rightmost violins represent the overall discrepancy between modelled treatment effects and modelled conditional treatment effects. Recall that this observed discrepancy in treatment effects, reclaimed in the true treatment effects, supports highly heterogenous treatment responsiveness and distributed mode of causation. SNRI=serotonin reuptake inhibitors; TCA=tricyclic antidepressants

**Table 2.** Treatment effects. The table shows for each class of migraine prophylactics the out-of-sample modelled average treatment effect; the out-of-sample modelled conditional average treatment effect; the out-of-sample overall and conditional true treatment effect (true response rates); and the accuracy of a logistic regression fitting the modelled individualized treatment effects to true response. Although the magnitude of the absolute increase in true treatment responders in the conditional subgroups may seem small, viewing these figures in light of the already small treatment effects highlights the significance of the difference. Moreover, the high accuracy of the regression models validates the generalizability of the findings.

| Intervention class (n) | Modelled average treatment effect (interquartile range) | Modelled conditional average treatment effect (interquartile range) | Overall true treatment effect (overall response rate) | Conditional true treatment effect (response rate in conditional subgroup) | Accuracy of 10-fold cross-validated logistic regression of predictions compared to true outcomes (standard deviation) |
| --- | --- | --- | --- | --- | --- |
| Botulinum toxin (111) | 0.44 (0.33-0.56) | 0.56 (0.48-0.65) | 0.47 | 0.51 | 0.53 (0.03) |
| Flunarizine (42) | 0.31 (0.23-0.42) | 0.42 (0.35-0.49) | 0.64 | 0.76 | 0.64 (0.11) |
| Candesartan (25) | 0.27 (0.15-0.41) | 0.42 (0.36-0.47) | 0.16 | 0.17 | 0.87 (0.16) |
| Tricyclic antidepressants (185) | 0.22 (0.14-0.32) | 0.32 (0.25-0.41) | 0.31 | 0.35 | 0.69 (0.02) |
| Valproate (68) | 0.17 (0.14-0.32) | 0.29 (0.20-0.36) | 0.26 | 0.32 | 0.74 (0.05) |
| Topiramate (121) | 0.16 (0.10-0.27) | 0.27 (0.20-0.33) | 0.29 | 0.28 | 0.71 (0.04) |
| Serotonin noradrenalie uptake inhibitors (66) | 0.16 (0.13-0.22) | 0.22 (0.20-0.30) | 0.29 | 0.33 | 0.71 (0.05) |
| Acupuncture (74) | 0.13 (0.10-0.19) | 0.19 (0.16-0.28) | 0.28 | 0.24 | 0.72 (0.03) |
| Betablockers (146) | 0.11 (0.06-0.18) | 0.18 (0.14-0.34) | 0.14 | 0.16 | 0.86 (0.02) |
| Serotonergic (55) | 0.06 (0.04-0.09) | 0.09 (0.08-0.11) | 0.16 | 0.22 | 0.84 (0.06) |

### Machine prescription and its impact

Out of the 253 patients included in the test set, 85 (33.6%) had tried their model predicted best treatment, 140 (55.3%) patients had tried at least one of their top two treatments, and 170 (67.2%) had tried at least one of the top three treatments.

Sequentially evaluating treatments by machine prescription resulted in arriving at a successful treatment in significantly fewer months than administering treatments in order by generic guideline recommendations (−3.750 months; 95% CI -3.993 to - 3.507; p<0.0001), or indeed any other justifiable order, including experienced clinician rankings (Figure 2 and Supplementary Table 5). The best treatment policy did not differ from randomly evaluating therapies. Finally, the average additional three-monthly cost for machine prescription was -£2 for low drug tariff estimates and £1 for high drug tariff estimates.

In the sensitivity analyses, reduction in time-to-response was –3.782 months (95% CI -4.574 to -2.990; p<0.0001) in the high-severity strata; and –3.343 months (95% CI - 4.860 to -1.825, p<0.0001) in the low-severity strata (Table 3).

**Figure 2.**
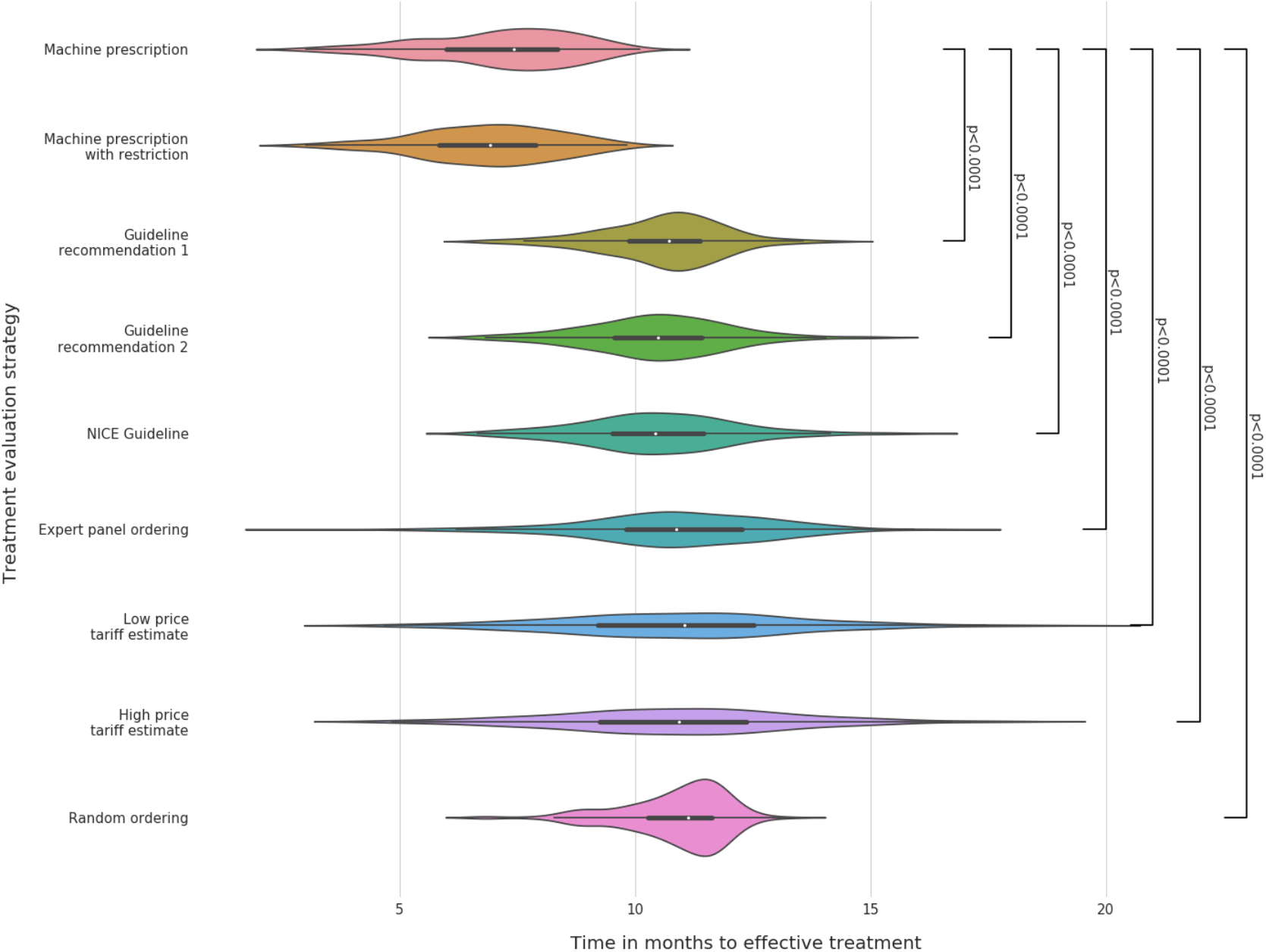
Time-to-response. Time-to-response given different strategies to decide sequence of evaluating treatments.

**Table 3.** Sensitivity analysis.

| **Analysis** | **Severely affected patients** | **Less severely affected patients** |
| --- | --- | --- |
| Definition | 25 or more headache days/month AND headache exacerbation severity of $\geq 9$ (on a VAS scale 0-10). | Fewer than 25 headache days/month AND headache exacerbation severity of $< 9$ (on a VAS scale 0-10). |
| Time-to-response with restricted machine prescription, mean (standard deviation) | 6.09 (2.06) | 3.52 (2.36) |
| Time-to-response with Guideline recommended sequence 1, mean (standard deviation) | 9.90 (2.49) | 7.14 (4.14) |
| Restricted ML model vs. guideline recommended ordering 1 | -3.782 months (95% confidence interval -4.574 to -2.990; $p < 0.0001$) | -3.343 months (95% confidence interval -4.860 to -1.825, $p < 0.0001$) |
| Additional cost with machine prescription, low drug tariff | -£57 | -£94 |
| Additional cost with machine prescription, high drug tariff | £18 | £2 |

## Discussion

Surveying a chronic migraine population amongst the largest and most finely phenotyped in the literature, here we show treatment heterogeneity to be robustly predictable from high-dimensional causal modelling of routinely collected clinical data. This finding supports a complex, distributed underlying mode of causation in chronic migraine, and suggests that neither the pursuit of a unitary causal mechanism, nor the evaluation of treatment effects within conventional randomised controlled trials is likely to be productive. Rather, deeper characterisation of patient heterogeneity is likely to be needed, through modelling richer additional features, such as imaging, physiological and genetic data^12-18^ at larger data scales, illuminating the wide causal field of factors that clearly underpins this complex disorder.

We show further that current treatment policy guidelines yield broadly the same time-to-response as chance. This is consistent with the widespread belief amongst clinicians that the individual selection of optimal treatment based on a small subset of individual patient factors is very difficult,^6^ a belief reinforced by expert panel rankings of treatments (Supplementary Table 3). By contrast, machine prescription offers a significantly shorter time-to-response, with a substantial mean effect size exceeding three months—equating to a 35% reduction. Crucially, better treatment is here achieved without a marked increase in cost, or plausibly greater risk of side effects,^19,20^ and without substantial variability across different severity strata. Close consideration must clearly be given to adopting the approach at scale, for the balance of risks and benefits is here heavily weighted in our favour.

It may seem premature to draw so general a set of conclusions from a single centre, tertiary referral population, even if this is one of the largest reported in the literature.

But it is crucial to appreciate that all inferences are here drawn from out-of-sample test data, indicating generalisability beyond the training data. Moreover, if a marked discrepancy between individual and population responsiveness is robustly identified within a comparatively small population with lower levels of heterogeneity than are observed in wider care, *a larger scale analysis can only magnify it*. This is because the tractability of patterns of heterogeneity can only be enhanced with data of greater scale and inclusivity. Indeed, our analysis invites replication with primary care data which we here show can be readily performed automatically with structured clinical records. The objective of this study is less to derive a set of specific models than to illustrate the optimal way of approaching machine prescription in migraine, given its manifest complexity.

Though a small proportion of patients did not fulfil strict diagnostic criteria, they were judged by an expert headache specialist to have chronic migraine, and our population demographics overall are in line with other large chronic migraine cohorts.^21,22^ Such patients should, and generally would, be treated as chronic migraine in real-world practice, and excluding them would limit rather than enhance generalisability to the wider population. Without objectively determinable aetiological criteria, all classification is in any event heuristic. That the conclusions here apply to a slightly broader population does not make them invalid: it extends their reach.

Finally, though allocation bias inevitably corrupts observational studies, in the context of heterogeneous treatment effects clearly exemplified here, it is only *one* contributory factor to the fidelity of the inference, and becomes increasingly *secondary* to the quality of the outcome model as the scale of available data rises.^23^ In any event, a multi-agent randomized controlled trial is obviously infeasible here, and evaluated with conventional statistics would be critically confounded by the individual-level, high-dimensional patterns of treatment responsiveness we have already demonstrated.

## Conclusion

Our analysis of a large and richly characterized dataset of chronic migraine phenotypes demonstrates—not only the value—but arguably the necessity of high-dimensional modelling in the management of migraine. We develop and evaluate a causal model of commonly used anti-migraine preventives that demonstrate both the distributed mode of causation, and the feasibility of machine prescription at the individual level. We conclude that the application of high-dimensional modelling to prescribing is a critical step towards reducing the massive global burden of migraine through realizing the personalized, precision medicine this remarkably complex condition demands.

## Acknowledgements

The authors have no acknowledgements.

## Author Contributions

AS: study concept, study design, data collection, data management, data analysis, interpretation of data, drafting and revision of manuscript.

RG: data analysis, interpretation of data, and manuscript revision.

ET: study concept, interpretation of data and manuscript revision.

MSM: study concept, study design, data collection, interpretation of data and manuscript revision.

PN: study concept, study design, data analysis, interpretation of data and manuscript revision.

## Abbreviated Summary

Stubberud et al. report that high dimensional modelling of the vastly heterogenous symptomatology of chronic migraine may predict optimal preventative treatment at the individual level. Using state-of-the-art machine learning they demonstrate that time-to-response is reduced by 35% compared to population-based policies.

